# Lack of weight gain and increased mortality during and after treatment among adults with drug-resistant tuberculosis in Georgia, 2009-2020

**DOI:** 10.1101/2024.08.05.24311499

**Authors:** Tsira Chakhaia, Henry M Blumberg, Russell R Kempker, Ruiyan Luo, Nino Dzidzikashvili, Mamuka Chincharauli, Nestan Tukvadze, Zaza Avaliani, Christine Stauber, Matthew J. Magee

## Abstract

**Background:** While low body mass index (BMI) is associated with poor tuberculosis (TB) treatment outcomes, the impact of weight gain during TB treatment is unclear. To address this knowledge gap, we assessed if lack of weight gain is associated with all-cause mortality during and after TB treatment.

**Methods:** We conducted a retrospective cohort study among adults with newly diagnosed multi- or extensively drug-resistant (M/XDR) pulmonary TB in Georgia between 2009-2020. The exposure was a change in BMI during the first 3-6 months of TB treatment. All-cause mortality during and after TB treatment was assessed using the National Death Registry. We used competing-risk Cox proportional hazard models to estimate adjusted hazard ratios (aHR) between BMI change and all-cause mortality.

**Results:** Among 720 adult participants, 21% had low BMI (<18.5 kg/m^2^) at treatment initiation and 9% died either during (n=16) or after treatment (n=50). During the first 3-6 months of TB treatment, 17% lost weight and 14% had no weight change. Among 479 adults with normal baseline BMI ( ≥18.5–24.9 kg/m^2^), weight loss was associated with an increased risk of death during TB treatment (aHR=5.25; 95%CI: 1.31-21.10). Among 149 adults with a low baseline BMI, no change in BMI was associated with increased post-TB treatment mortality (aHR=4.99; 95%CI: 1.25-19.94).

**Conclusions:** Weight loss during TB treatment (among those with normal baseline BMI) or no weight gain (among those with low baseline BMI) was associated with increased rates of all-cause mortality. Our findings suggest that scaling up weight management interventions among those with M/XDR TB may be beneficial.

**Summary:** Among a cohort of persons with drug resistant tuberculosis (TB), failure to gain weight during TB treatment was associated with an increased risk of all-cause mortality during and after completion of treatment.

## Introduction

Low body mass index (BMI) is an established risk factor for tuberculosis (TB) and predicts worse TB treatment outcomes including higher mortality rates (1–7). Prior studies also describe a dose-response relationship with low BMI at TB treatment initiation and poor TB treatment outcomes (8–15). However, it is unclear whether lack of weight gain during TB treatment exacerbates the deleterious effects of low baseline BMI on treatment outcomes among patients with TB. Some studies suggest that weight gain during treatment may be associated with favorable treatment outcomes (16, 17). Early evidence suggests that weight gain during TB treatment may have a particular benefit among those who are underweight at the start of treatment (18). Additional data are needed to understand the relationship between extent of weight change during TB treatment and TB treatment outcomes, including the impact of weight change on mortality during and after TB.

Given the current knowledge gaps in the understanding of the impact of weight change during TB treatment on treatment outcomes, the overall goal of our study was to assess the relationship between BMI change during treatment and TB treatment outcomes among persons with multi- or extensively drug-resistant (M/XDR) TB. We estimated the association between the change in BMI with all-cause mortality during treatment and post-TB. We also examined whether the primary associations differed by BMI at TB treatment initiation

## Methods

### Study Design and Settings

We conducted a cohort study using secondary data from the Georgia National TB Program (NTP). Acid fast bacillus (AFB) sputum smear, Gene-Xpert and XpertMTB/XDR, line probe assay (LPA), culture, and drug susceptibility testing (DST), chest X-ray, complete blood counts and fasting blood glucose (FBG) tests, weight and height measurements were conducted among patients with M/XDR-TB before treatment initiation. Some of these tests were also performed at monthly visits during treatment. In Georgia, persons with confirmed M/XDR TB receive ambulatory care treatment or are initially hospitalized based on the patients’ clinical status. All treatment is provided by directly observed therapy (DOT). Although Georgia first utilized the new anti-TB drug bedaquiline through compassionate use from 2011 to 2014, and later as part of a new DR-TB treatment program supported by MSF-France from 2014 to 2016, it was not until 2016 that the Georgian NTP formally incorporated new and repurposed anti-TB drugs such as bedaquiline, delamanid, and linezolid for the treatment of M/XDR-TB (19, 20).

### Study population

Persons ≥16 years old with new, bacteriologically confirmed M/XDR pulmonary TB registered in Georgian TB facilities during 2009-2020 who had completed TB treatment by the end of 2021 were eligible for inclusion in our study. Patients who did not have baseline weight and height measurements or follow up weight measurements during TB treatment were excluded. We excluded persons from the final analysis if they had their weight monitored only before the third month or after the sixth month of treatment, or if they died or discontinued treatment before the third month. This allowed us to assess the change in BMI from the start of M/XDR-TB treatment to the first 3-6 months of treatment.

### Data sources and data collection

We obtained data on participants’ demographic information and socioeconomic and clinical characteristics, including the data on weight change during TB treatment, from medical charts and the Georgian NTP electronic database. Additionally, the Georgia National Death Registry was used to link patient information with vital status after TB treatment. The vital status of the participants was queried on 28 February 2023. We linked the different data sources using patient name, date of birth, unique code assigned to each patient upon registration in the Georgian NTP, and/or national identification number. All collected data were entered into an online Research Electronic Data Capture (REDCap) database hosted at Georgia State University (21).

### Data variables and definitions

The primary exposure of interest for this study was change in BMI from the start of M/XDR-TB treatment to the first 3-6 months of treatment categorized into negative change, no change, and positive change. If a patient had >1 weight measurements between the third and sixth months of treatment, the nearest measurement to the 3^rd^ month was chosen for calculation of change in BMI. We also calculated relative change in BMI, defined as the percentage change from the start of treatment to the first 3-6 months of treatment and a rate of relative BMI change per three person months of treatment. The primary outcome of interest was all-cause mortality during TB treatment, defined as deaths during the current TB treatment episode and registered in the NTP electronic database. The secondary outcome of interest was all-cause post-TB mortality, defined as deaths registered in the National Death Registry and dated between the date of TB treatment outcome and the 28^th^ of February 2023 when queries on mortality data were requested.

We categorized the patients as underweight/low BMI (BMI < 18.5 kg/m^2^), normal weight/normal BMI (BMI: 18.5 - 25.0 kg/m^2^), and overweight or obese/high BMI (BMI ≥ 25.0 kg/m^2^) (22). Covariates included age, gender, smoking status, drug resistance profile, HIV and HCV serologic status, AFB sputum smear status at baseline, and year of MDR-TB treatment initiation.

### Statistical analysis

We compared patient characteristics across the categories of negative, no, and positive change in BMI using chi-square or Fisher’s exact test for categorical predictors, and t-test or Wilcoxon rank sum for continuous variables. Sociodemographic, behavioral, and clinical characteristics identified by directed acyclic graphs (DAG) or with bivariate p-value ≤ 0.05 (with both exposure and outcome) were included in the multivariable analysis, and adjusted hazard rate ratios (aHRs) with 95% confidence intervals (CIs) were estimated with competing risk Cox regression. For the outcome – death during TB treatment, patients who did not die during TB treatment were censored on the date of their TB treatment outcome. For the outcome – death after completion of TB treatment, participants were censored 1) at the death date if they died during treatment (competing risk), 2) on 28 February 2023 if they did not die until the date mortality status was obtained; or 3) at the day of their TB treatment outcome date, if they were not listed as dead in the Georgian National Death Registry. We assessed the proportional hazard assumptions with Schoenfeld’s residuals tests (23). In addition, we examined the unadjusted and adjusted associations of change in BMI with outcomes in the cohorts stratified by baseline BMI categories (low, normal, and high BMI) and MDR-TB treatment initiation period (2016-2020 when new anti-TB drugs were used vs 2009-2015 during a period before the introduction of new anti-TB drugs).

### Sensitivity analysis

Because a substantial proportion of (34.8%) eligible patients did not have available data on change in BMI throughout the first 3-6 months of treatment (and were not included in the final analyses), we compared baseline characteristics of the cohort included in the study vs. those excluded from the study to determine any differences between these two groups. In addition, we estimated the cumulative hazard rates for all-cause mortality during treatment in the included and excluded cohorts.

### Ethical considerations

The study was approved by the Institutional Review Boards (IRBs) at the National Center for Tuberculosis and Lung Disease (Tbilisi, Georgia) and at Georgia State University (Atlanta, GA, USA).

## Results

### Baseline characteristics of M/XDR TB patients

A total of 720 eligible study participants with M/XDR TB who had baseline and follow up BMI measures were included (**Figure 1**). The median age of participants was 35.5 years (IQR: 26.5-49.0), and 68.8% were male. Most participants had positive AFB sputum smear microscopy (63.5%) results at MDR-TB treatment initiation. During a period when there was the availability of new and repurposed anti-TB drugs (2016-2020), 329 (45.7%) were enrolled in MDR-TB treatment. Among study participants, 149 (20.7%) had low baseline BMI, and 92 (12.8%) were either overweight or obese at MDR-TB treatment initiation. Median baseline BMI was 20.6 kg/m^2^ (IQR: 18.8-23.0). Among all patients, 123 (17.1%) had a decrease in BMI, and 499 (69.3%) had an increase in BMI during the first 3-6 months of MDR-TB treatment **(Table 1)**. The median relative change in BMI during these initial months was an increase of 2.8% (IQR: 0%-6.3%). Among those who lost or gained weight, the median baseline BMI was 20.8 kg/m^2^ (IQR: 18.9-24.6) and 20.5 kg/m^2^ (IQR: 18.8-22.7), respectively (**Table 1).** Overall, 46 (30.9%) of 149 with a low baseline BMI and 135 (28.2%) of 479 with normal baseline BMI either lost or did not gain weight during the first three-six months of TB treatment. More than half (56.5%) of overweight/obese patients had positive change in BMI **(Table 1)**.

**Figure 1.**
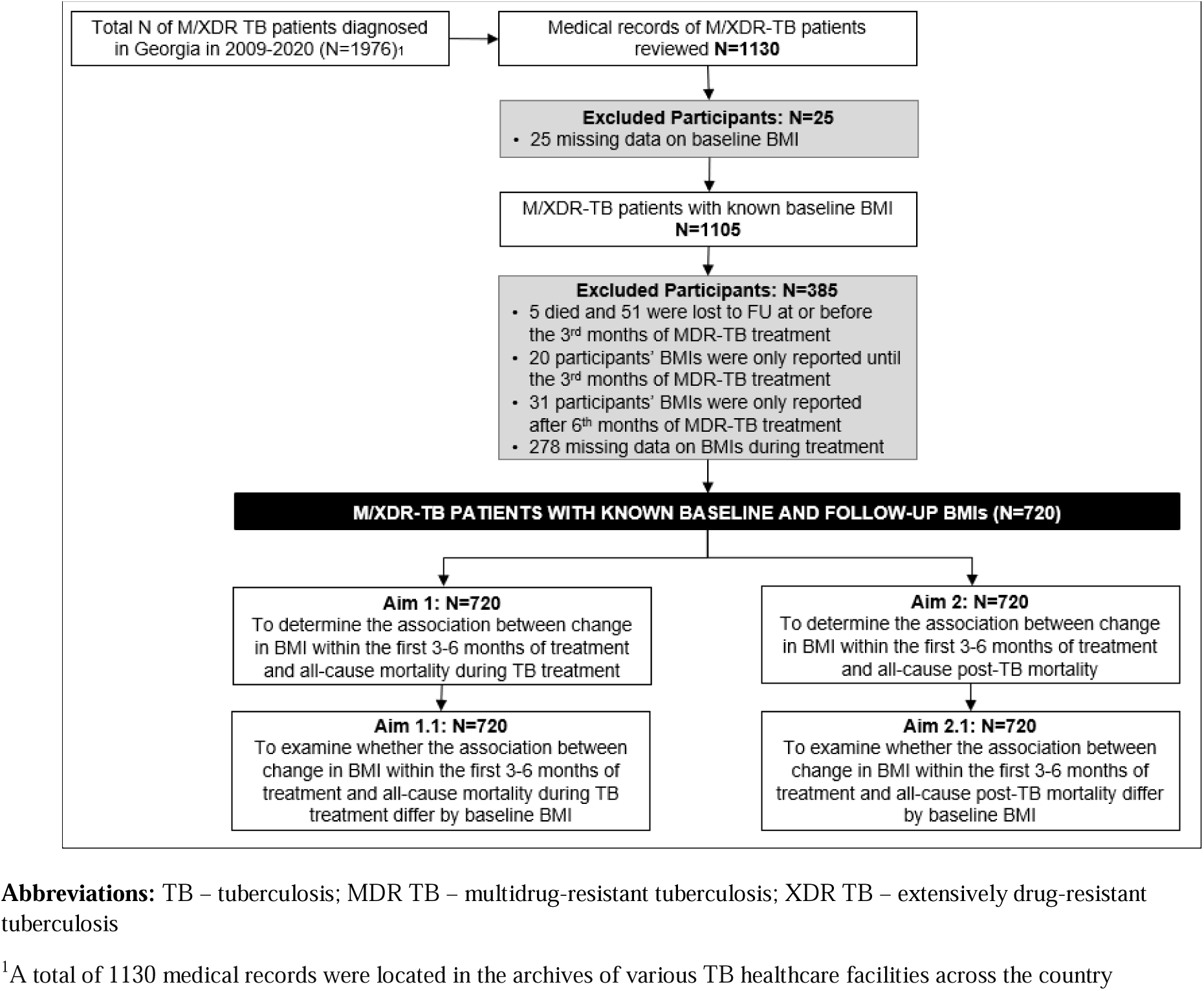
Flowchart of study participants with M/XDR-TB, 2009-2020, Georgia.

**Table 1.**
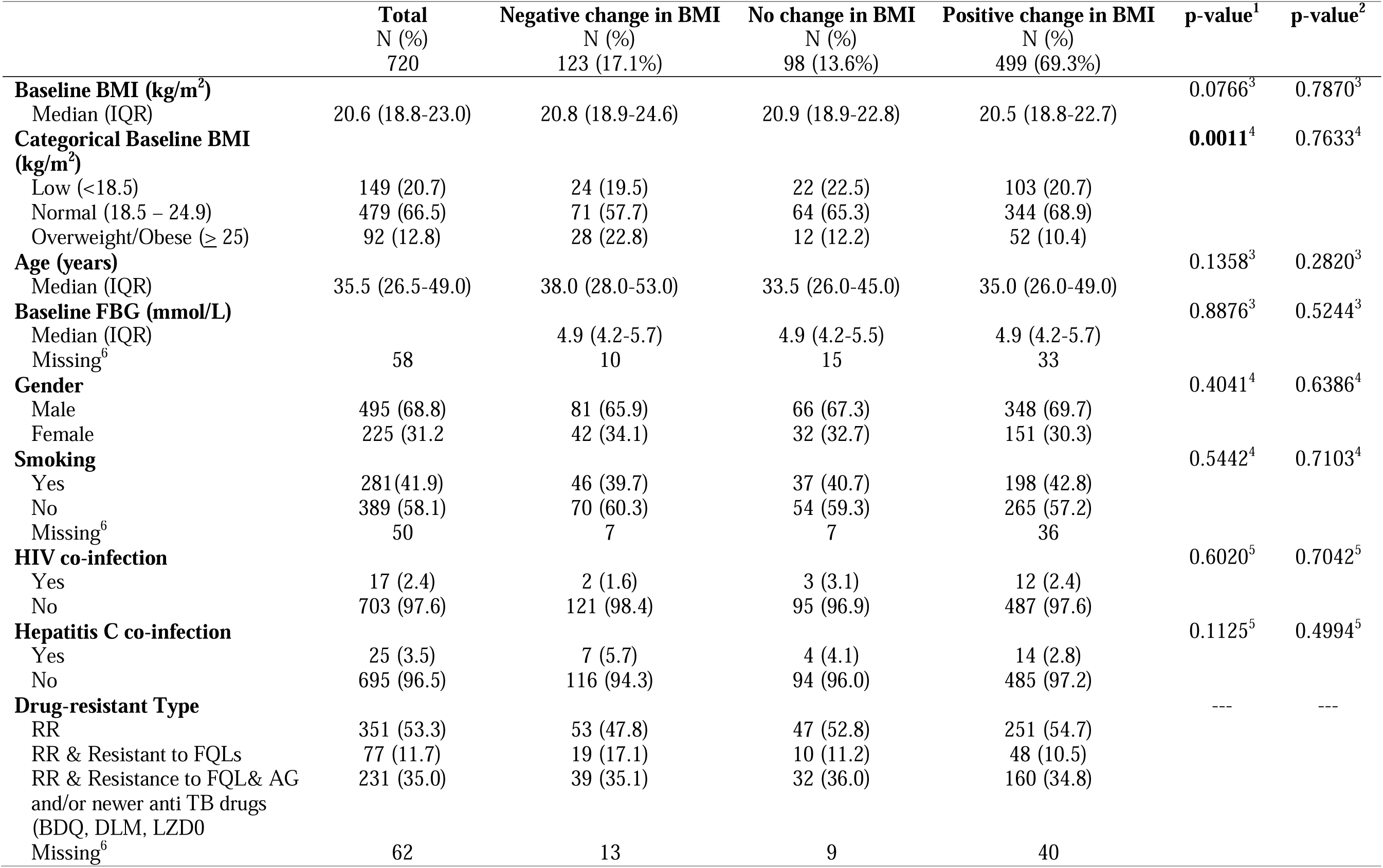
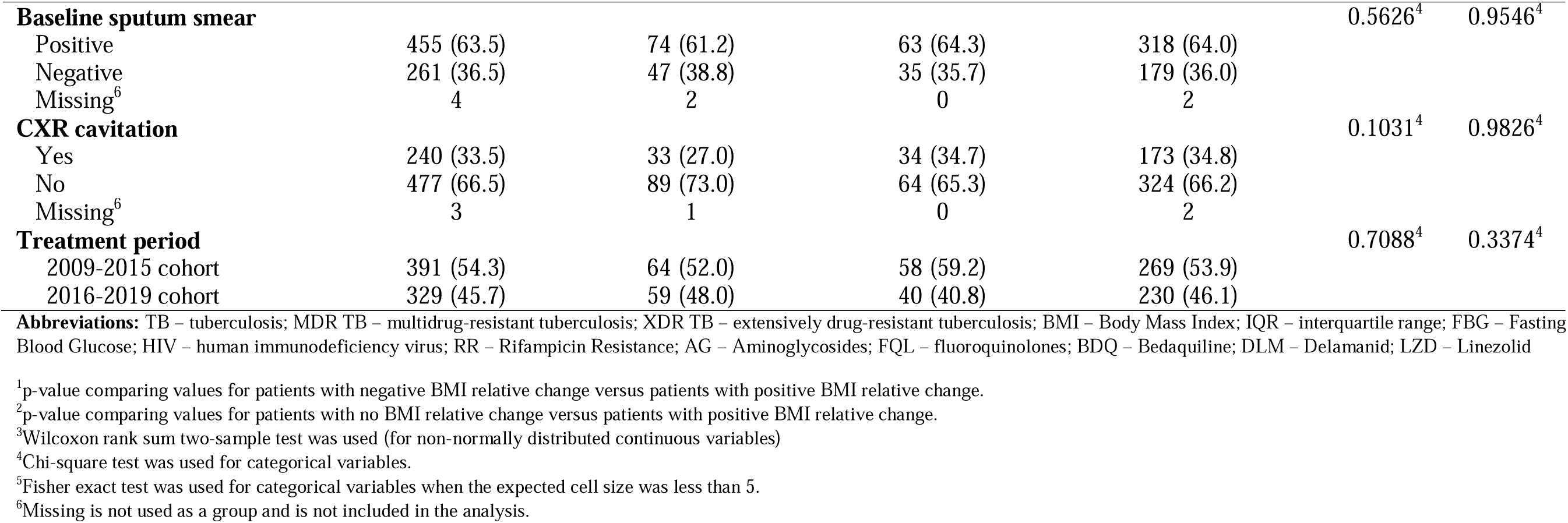
Patient and clinical baseline characteristics among M/XDR TB patients by change in BMI, Georgia, 2009-2020 (N=720)

### Association between change in BMI and all-cause mortality during and after TB treatment

Among 720 patients with M/XDR TB, 16 (2.2%) died during and 50 (6.9%) died after TB treatment (Table 2). Those with older age (HR=1.04 per month; 95% CI: 1.01-1.08) and those with HIV infection (HR=7.81; 95% CI: 1.76-34.67) were at increased risk for death during TB treatment. The cumulative incidence of death during treatment was 4.1% among those with weight loss, 3.1% in those with no change, and 1.6% in those with positive BMI change. **Supplemental Figure A** presents mortality during and after TB treatment by change in BMI during MDR-TB treatment.

**Table 2.**
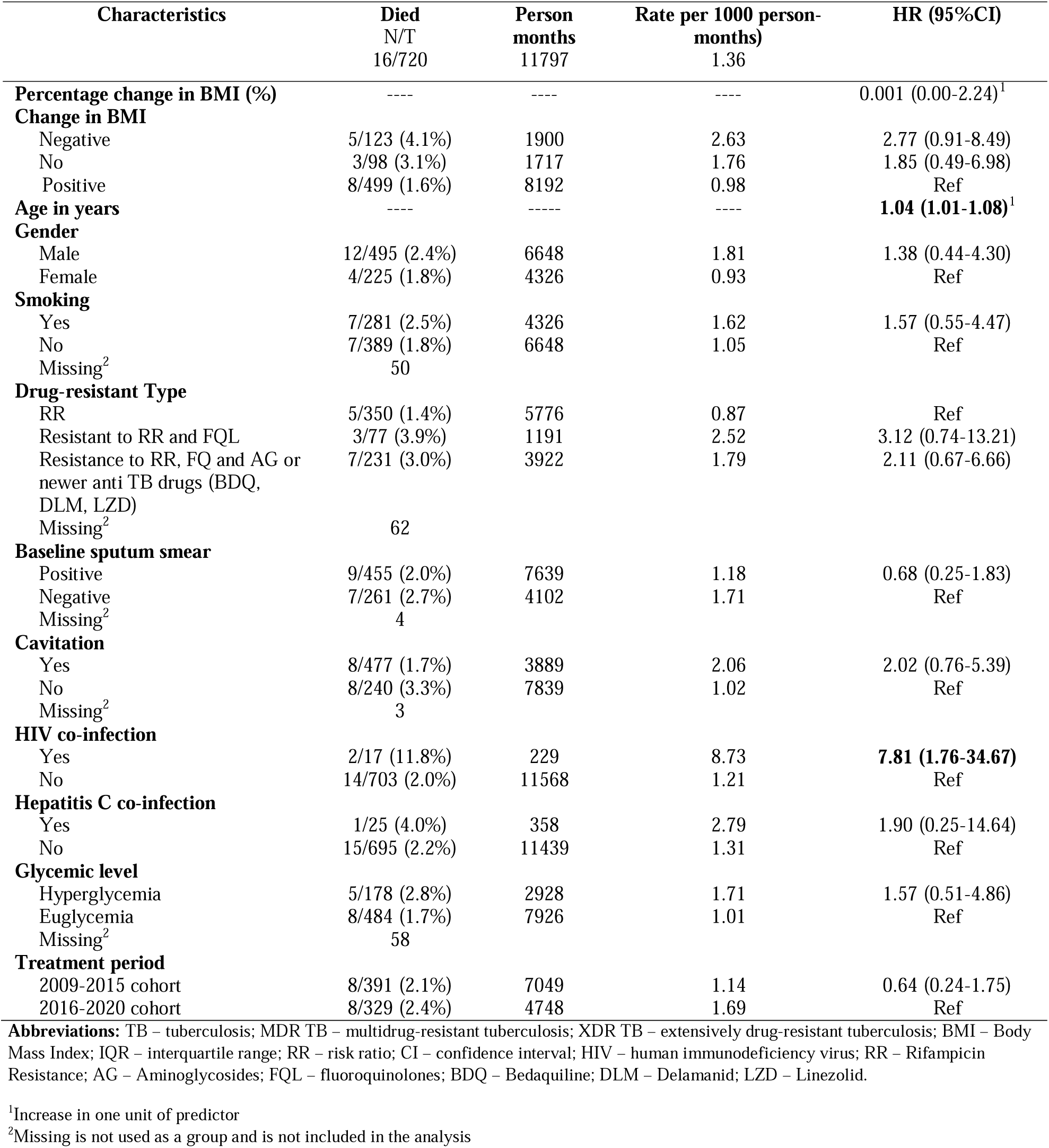
Demographic and clinical characteristics at MDR-TB treatment enrollment associated with death during TB treatment among M/XDR TB patients in Georgia, 2009-2020 (N=720)

| Characteristics | Died<br>N/T<br>16/720 | Person<br>months<br>11797 | Rate per 1000 person-<br>months)<br>1.36 | HR (95%CI) |
| --- | --- | --- | --- | --- |
| **Percentage change in BMI (%)** | ---- | ---- | ---- | 0.001 (0.00-2.24)<sup>1</sup> |
| **Change in BMI** |  |  |  |  |
| Negative | 5/123 (4.1%) | 1900 | 2.63 | 2.77 (0.91-8.49) |
| No | 3/98 (3.1%) | 1717 | 1.76 | 1.85 (0.49-6.98) |
| Positive | 8/499 (1.6%) | 8192 | 0.98 | Ref |
| **Age in years** | ---- | ----- | ---- | **1.04 (1.01-1.08)<sup>1</sup>** |
| **Gender** |  |  |  |  |
| Male | 12/495 (2.4%) | 6648 | 1.81 | 1.38 (0.44-4.30) |
| Female | 4/225 (1.8%) | 4326 | 0.93 | Ref |
| **Smoking** |  |  |  |  |
| Yes | 7/281 (2.5%) | 4326 | 1.62 | 1.57 (0.55-4.47) |
| No | 7/389 (1.8%) | 6648 | 1.05 | Ref |
| Missing<sup>2</sup> | 50 |  |  |  |
| **Drug-resistant Type** |  |  |  |  |
| RR | 5/350 (1.4%) | 5776 | 0.87 | Ref |
| Resistant to RR and FQL | 3/77 (3.9%) | 1191 | 2.52 | 3.12 (0.74-13.21) |
| Resistance to RR, FQ and AG or newer anti TB drugs (BDQ, DLM, LZD) | 7/231 (3.0%) | 3922 | 1.79 | 2.11 (0.67-6.66) |
| Missing<sup>2</sup> | 62 |  |  |  |
| **Baseline sputum smear** |  |  |  |  |
| Positive | 9/455 (2.0%) | 7639 | 1.18 | 0.68 (0.25-1.83) |
| Negative | 7/261 (2.7%) | 4102 | 1.71 | Ref |
| Missing<sup>2</sup> | 4 |  |  |  |
| **Cavitation** |  |  |  |  |
| Yes | 8/477 (1.7%) | 3889 | 2.06 | 2.02 (0.76-5.39) |
| No | 8/240 (3.3%) | 7839 | 1.02 | Ref |
| Missing<sup>2</sup> | 3 |  |  |  |
| **HIV co-infection** |  |  |  |  |
| Yes | 2/17 (11.8%) | 229 | 8.73 | **7.81 (1.76-34.67)** |
| No | 14/703 (2.0%) | 11568 | 1.21 | Ref |
| **Hepatitis C co-infection** |  |  |  |  |
| Yes | 1/25 (4.0%) | 358 | 2.79 | 1.90 (0.25-14.64) |
| No | 15/695 (2.2%) | 11439 | 1.31 | Ref |
| **Glycemic level** |  |  |  |  |
| Hyperglycemia | 5/178 (2.8%) | 2928 | 1.71 | 1.57 (0.51-4.86) |
| Euglycemia | 8/484 (1.7%) | 7926 | 1.01 | Ref |
| Missing<sup>2</sup> | 58 |  |  |  |
| **Treatment period** |  |  |  |  |
| 2009-2015 cohort | 8/391 (2.1%) | 7049 | 1.14 | 0.64 (0.24-1.75) |
| 2016-2020 cohort | 8/329 (2.4%) | 4748 | 1.69 | Ref |

**Abbreviations:** TB – tuberculosis; MDR TB – multidrug-resistant tuberculosis; XDR TB – extensively drug-resistant tuberculosis; BMI – Body Mass Index; IQR – interquartile range; RR – risk ratio; CI – confidence interval; HIV – human immunodeficiency virus; RR – Rifampicin Resistance; AG – Aminoglycosides; FQL – fluoroquinolones; BDQ – Bedaquiline; DLM – Delamanid; LZD – Linezolid.
<sup>1</sup>Increase in one unit of predictor
<sup>2</sup>Missing is not used as a group and is not included in the analysis

In multivariate analysis, the aHR of all-cause mortality during TB treatment was 2.63 (95% CI: 0.85-8.08) for those with negative change in BMI compared to those with positive change in BMI. The aHR of all-cause mortality post-TB treatment was 1.99 (95% CI: 0.96-4.11) for those with no change compared to those with positive change in BMI. When participants were stratified by BMI at MDR-TB treatment initiation, among 149 study participants with low BMI at TB treatment initiation, those who had no change in BMI (compared to those with a positive change in BMI) were at increased risk of all-cause post-TB mortality (HR=4.42; 95% CI: 1.18 – 16.47). After adjusting for age, gender, year of treatment initiation, baseline sputum smear results and cavitation on chest radiograph, the association remained significant (aHR=4.99; 95% CI: 1.25-19.94). Among 479 adults with normal BMI at treatment initiation, weight loss compared to weight gain was statistically significantly associated with an increased risk of death during TB treatment in unadjusted (HR = 5.11; 95% CI: 1.28-20.44) and adjusted models (aHR = 5.25; 95%CI: 1.31-21.10) **(Table 3)**.

**Table 3.** Competing risk hazard rates of all-cause mortality during and after treatment stratified by BMI at treatment initiation among adults with M/XDR TB patients, Georgia, 2009-2020 (N=720)

| Cohort | Change in BMI (n) | Cause-Specific Hazard Model<br>Hazard ratio (95% CI) |  |  |  |
| --- | --- | --- | --- | --- | --- |
|  |  | All-cause mortality during treatment<sup>1</sup> | All-cause mortality during treatment<sup>2</sup> | All-cause post-TB mortality<sup>1</sup> | All-cause post-TB mortality<sup>2</sup> |
| Total cohort (n = 720)<sup>1</sup> | Negative (123) | 2.61 (0.85 - 7.96) | 2.63 (0.85 - 8.08) | 1.44 (0.71 - 2.96) | 1.07 (0.51 - 2.22) |
|  | No (98) | 1.91 (0.51 - 7.21) | 2.10 (0.55 - 7.96) | 1.74 (0.85 - 3.56) | 1.99 (0.96 - 4.11) |
|  | Positive (499) | REF | REF | REF | REF |
| Cohort with baseline low BMI (<18.5 kg/m<sup>2</sup>) (n = 149) | Negative (24) | ----<sup>3</sup> | ----<sup>3</sup> | 2.75 (0.65 - 11.56) | 2.30 (0.52 - 10.22) |
|  | No (22) | 1.17 (0.13 - 10.48) | 2.69 (0.22 - 33.34) | **4.42 (1.18 - 16.47)** | **4.99 (1.25 - 19.94)** |
|  | Positive (103) | REF | REF | REF | REF |
| Cohort with baseline normal BMI (18.5-24.9 kg/m<sup>2</sup>) (n = 479) | Negative (71) | **5.11 (1.28 - 20.44)** | **5.25 (1.31 - 21.10)** | 1.09 (0.37 - 3.20) | 1.03 (0.35 - 3.03) |
|  | No (64) | 1.39 (0.16 - 12.47) | 1.26 (0.14 - 11.41) | 1.41 (0.53 - 3.77) | 1.56 (0.56 - 4.32) |
|  | Positive (344) | REF | REF | REF | REF |
| Cohort with baseline high BMI<sup>1</sup> (≥ 25 kg/m<sup>2</sup>) (n = 92) | Negative (28) | REF<sup>4</sup> | REF<sup>4</sup> | 0.93 (0.23 - 3.73) | **0.10 (0.01 - 0.94)** |
|  | No (12) | 2.13 (0.13 - 33.97) | 2.73 (0.01 - 34.99) | 0.62 (0.07 - 5.23) | 0.80 (0.07 - 9.40) |
|  | Positive (52) | ----<sup>3</sup> | ----<sup>3</sup> | REF | REF |

**Abbreviations:** TB – tuberculosis; MDR TB – multidrug-resistant tuberculosis; XDR TB – extensively drug-resistant tuberculosis; BMI – Body Mass Index; CI – confidence interval.
<sup>1</sup>Unadjusted
<sup>2</sup>Adjusted for gender, age, year of MDR-TB treatment initiation, baseline sputum smear and chest radiography cavitation.
<sup>3</sup>No death observed for this group.
<sup>4</sup>In the low and normal baseline BMI cohorts, the reference group consisted of individuals with a positive change in BMI. In the high baseline BMI cohort, no deaths occurred during treatment among those with a positive change in BMI. Therefore, to evaluate mortality during treatment, the reference group comprised individuals with a negative change in BMI.

### Sensitiivty Analysis: Baseline characteristics and mortality rate among included and excluded participants

Baseline characteristics were similar between participants included and excluded from the study, except for age, baseline sputum smear results, and treatment period. The median age of the included and excluded individuals was 35.5 (IQR: 26.5-49.0) and 34.0 (IQR: 25.0-46.0) (p-value=0.03) respectively. More participants in the included cohort had positive sputum smear results at treatment initiation (63.5% vs. 51.0%; p-value<0.01). In addition, persons excluded from the study started MDR-TB treatment in 2009-2015 (65.2%) compared to 54.3% of those included in the study cohort (p-value<0.01) **(Table 4)**. Difference in mortality during treatment between included and excluded participants was not statistically significant **(Supplemental Table B)**.

**Table 4.**
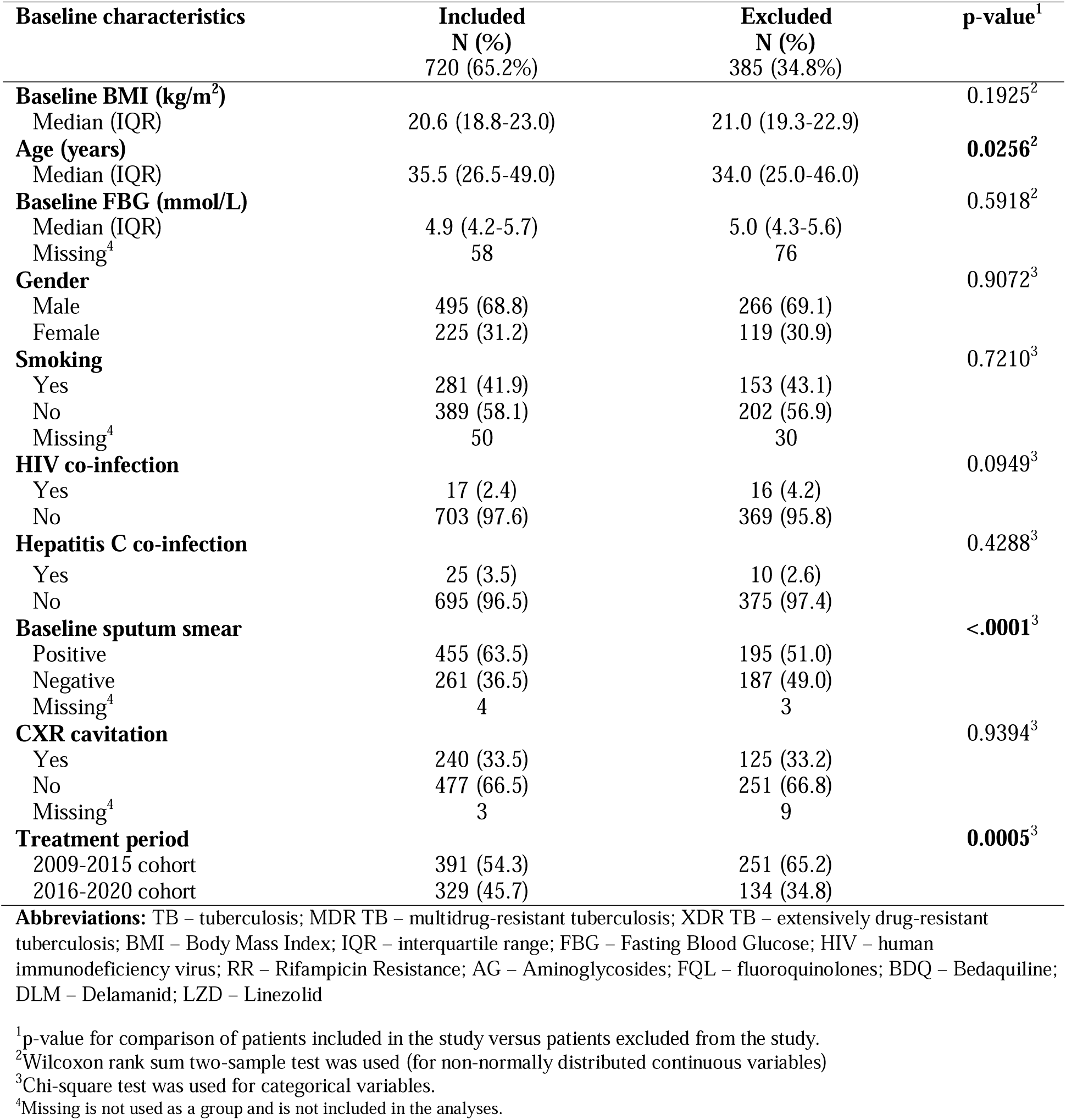
Patient and clinical baseline characteristics among M/XDR TB patients included in and excluded from the study, Tbilisi, Georgia, 2009-2020 (N=1105)

| Baseline characteristics | Included<br>N (%)<br>720 (65.2%) | Excluded<br>N (%)<br>385 (34.8%) | p-value<sup>1</sup> |
| --- | --- | --- | --- |
| **Baseline BMI (kg/m<sup>2</sup>)** |  |  | 0.1925<sup>2</sup> |
| Median (IQR) | 20.6 (18.8-23.0) | 21.0 (19.3-22.9) |  |
| **Age (years)** |  |  | **0.0256<sup>2</sup>** |
| Median (IQR) | 35.5 (26.5-49.0) | 34.0 (25.0-46.0) |  |
| **Baseline FBG (mmol/L)** |  |  | 0.5918<sup>2</sup> |
| Median (IQR) | 4.9 (4.2-5.7) | 5.0 (4.3-5.6) |  |
| Missing<sup>4</sup> | 58 | 76 |  |
| **Gender** |  |  | 0.9072<sup>3</sup> |
| Male | 495 (68.8) | 266 (69.1) |  |
| Female | 225 (31.2) | 119 (30.9) |  |
| **Smoking** |  |  | 0.7210<sup>3</sup> |
| Yes | 281 (41.9) | 153 (43.1) |  |
| No | 389 (58.1) | 202 (56.9) |  |
| Missing<sup>4</sup> | 50 | 30 |  |
| **HIV co-infection** |  |  | 0.0949<sup>3</sup> |
| Yes | 17 (2.4) | 16 (4.2) |  |
| No | 703 (97.6) | 369 (95.8) |  |
| **Hepatitis C co-infection** |  |  | 0.4288<sup>3</sup> |
| Yes | 25 (3.5) | 10 (2.6) |  |
| No | 695 (96.5) | 375 (97.4) |  |
| **Baseline sputum smear** |  |  | **<.0001<sup>3</sup>** |
| Positive | 455 (63.5) | 195 (51.0) |  |
| Negative | 261 (36.5) | 187 (49.0) |  |
| Missing<sup>4</sup> | 4 | 3 |  |
| **CXR cavitation** |  |  | 0.9394<sup>3</sup> |
| Yes | 240 (33.5) | 125 (33.2) |  |
| No | 477 (66.5) | 251 (66.8) |  |
| Missing<sup>4</sup> | 3 | 9 |  |
| **Treatment period** |  |  | **0.0005<sup>3</sup>** |
| 2009-2015 cohort | 391 (54.3) | 251 (65.2) |  |
| 2016-2020 cohort | 329 (45.7) | 134 (34.8) |  |

**Abbreviations:** TB – tuberculosis; MDR TB – multidrug-resistant tuberculosis; XDR TB – extensively drug-resistant tuberculosis; BMI – Body Mass Index; IQR – interquartile range; FBG – Fasting Blood Glucose; HIV – human immunodeficiency virus; RR – Rifampicin Resistance; AG – Aminoglycosides; FQL – fluoroquinolones; BDQ – Bedaquiline; DLM – Delamanid; LZD – Linezolid
<sup>1</sup>p-value for comparison of patients included in the study versus patients excluded from the study.
<sup>2</sup>Wilcoxon rank sum two-sample test was used (for non-normally distributed continuous variables)
<sup>3</sup>Chi-square test was used for categorical variables.
<sup>4</sup>Missing is not used as a group and is not included in the analyses.

## Discussion

Our study demonstrated that failure to gain weight during treatment for participants with M/XDR-TB was associated with an increased rate of all-cause mortality during and after TB treatment among those with low and normal BMI at the time of treatment initiation. Overall, we found that a high proportion (30.7%) of participants had lost (17.1%) or did not gain weight (13.6%) during the first 3-6 months of MDR-TB treatment. Patients with low baseline BMI who did not gain weight during the first 3-6 months of treatment had nearly five times the relative rate of all-cause post-TB mortality than those who gained weight. In addition, patients with normal baseline BMI who lost weight also had more than five times the relative rate of all-cause mortality during the treatment compared to those who gained weight. Thus, our study found an important association between failure to gain weight and weight loss during MDR-TB treatment and treatment outcome.

In our study, we obtained population-based data from more than ten years using multiple sources, including the Georgian NTP electronic database, paper-based medical charts, and the National Death Registry. The use of multiple population-based data sources helped ensure more complete and accurate data, which increases the rigor and generalizability of our findings. Additionally, the use of a comprehensive source of mortality data extended the scope of our study to include information on post-TB mortality and allowed us to assess the impact of BMI changes during TB treatment on long-term health outcomes.

The study presents stratified data indicating that a lack of weight gain during TB treatment amplifies the negative impact of low baseline BMI on treatment outcomes among patients with MDR-TB. Our results provide additional insights to prior studies including a 2016-2018 study from Guinea by Diallo et. al. among 165 patients with MDR-TB and a study by Chung-Delgado et. al. among 243 patients with MDR-TB from Peru. The prior studies from Guinea and Peru examined the correlation between weight change and TB treatment outcomes and found that the success of the MDR-TB treatment and the absence of lung cavities on the chest X-ray were associated with an increase in BMI in patients during TB treatment (16, 17). Unlike prior studies, our analyses specifically examined the impact of weight gain on mortality among patients with different baseline BMI levels. A study from Philippines by Gler et. al. among 439 patients with MDR-TB examined the association of weight gain with favorable TB treatment outcomes among underweight patients, and suggested that weight gain during TB treatment had a positive impact on treatment outcomes, but did not assess long-term outcomes such as post-TB mortality (18).

To our knowledge, our study is the first to assess the relationship between changes in BMI during TB treatment and treatment outcomes in Georgia. Previous studies in Georgia have reported on the relationship between baseline BMI and TB treatment outcomes as a secondary analysis, rather than as a primary focus of their research (8, 10, 12). Notably, a study by Adamashvili et al. in Georgia reported an association between BMI and all-cause post-TB mortality but did not specifically examine the role of BMI change during treatment (8).

Initial low BMI and lack of weight gain during TB treatment may be related to mortality through several biological pathways. First, TB is associated with a catabolic state which may induce or worsen malnutrition (24). Individuals with more severe TB are likely to experience wasting of lean and fat mass in addition to loss of micronutrients (25, 26). Therefore it is plausible that low BMI at the time of treatment initiation is a marker of more severe TB disease and consequently individuals with low BMI are also at increased risk of poor TB outcomes (27). Second, inadequate nutrition may accelerate *Mtb* growth, replication, and necrosis resulting in a chronic inflammatory response which can result in cavitation (28). Tissue damage from TB may disrupt glucose metabolism and further influence weight gain during TB treatment (29). Third, undernutrition may impact the absorption and metabolism of anti-TB drugs (30), which can increase the risk of TB treatment failure and mortality.

Addressing TB-related undernutrition issues requires comprehensive care, including weight monitoring and nutritional support. Our findings have critical implications for developing nutritional intervention studies among patients with TB. A randomized trial conducted in India concluded that nutritional intervention led to a substantial (39–48%) relative reduction in tuberculosis incidence among households contacts of index TB cases over a 2-year follow-up period. Given our study’s demonstration of the negative impact of weight loss during TB treatment on treatment outcomes, further investigations are needed to determine whether nutritional interventions are beneficial and improve treatment outcomes among those with active TB disease, particularly among individuals with low BMI during treatment (31).

Our study is subject to limitations. One-fourth of the eligible patients with M/XDR-TB did not have weight measurements reported during TB treatment, 5% did not have weight monitoring within the first 3-6 months of TB treatment, and an additional 5% either discontinued treatment or died before the third month of TB treatment and were excluded from the analyses. To address this limitation, we compared baseline characteristics of the excluded and included participants to determine if there were any significant differences that could potentially affect the study outcomes. Those excluded from the study were more likely to be diagnosed with TB in the earlier time period (2009-2015) compared to the more recent time period (2016-2020), (65.2% vs. 34.85) (p-value=<0.01). To ensure that the study findings are representative of the entire population and not biased due to missing data, we controlled the primary associations for the baseline characteristics by which included and excluded participants differed. In addition, analyzing cohorts enrolled in MDR-TB treatment in different years could result in biased findings because new and repurposed anti-TB drugs were introduced in 2016 for treatment of M/XDR-TB. Patient-level treatment regimen data was not included. To address this limitation, we analyzed participants stratified by time period of treatment initiation (before and after introducing new and repurposed TB drugs). Finally, this was a retrospective study. Thus, the data were not initially collected specifically for research purposes and there was some variability in the timing of weight monitoring during treatment. We addressed this issue of timing by calculating the rate of relative change in BMI per 90 days and including it as a continuous variable in the analysis.

## Conclusion

Our retrospective population-based cohort study that included persons with M/XDR-TB from Georgia demonstrated an association of weight gain with lower rates of all-cause mortality during and after TB treatment, especially among patients with low or normal baseline BMI. In analyses stratified by baseline BMI, weight loss during TB treatment (among those with normal BMI) or no weight gain (among those with low BMI) was associated with increased rates of all-cause mortality. Thus, the study’s findings suggest that weight gain during TB treatment is an important predictor of favorable treatment outcomes. These findings also have implications for future treatment guidelines and emphasize the need for studies on interventions for patients with drug-resistant (and drug susceptible) TB, especially those with undernutrition and assessing whether nutritional supplements can reduce mortality among patients at high risk for poor outcomes.

## Data Availability

All data produced in the present study are available upon reasonable request to the authors

## Acknowledgements

TC, HMB, RRK, CS, and MJM conceptualized the study design. TC, RL, and MJM led data analysis and interpretation of data in drafting the manuscript. TC, ND, MC, ZA and NT participated in key leadership of data collection. All provided critical revision of the article, interpretation of the data, and approved the final manuscript.

The authors have no conflicts of interest to declare.

## Supplementary Material

**Supplemental Table A.**
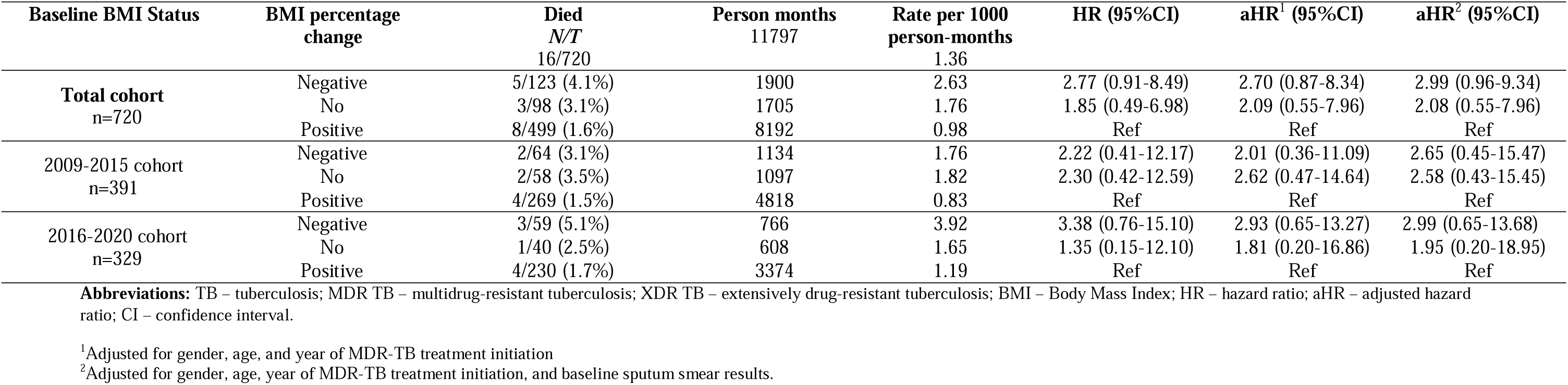
Associations between change in BMI with all-cause mortality during TB treatment stratified by period of MDR-TB treatment initiation, among adult M/XDR TB patients, Georgia, 2009-2020 (N=720). **Purpose:** To present the hazard of death during TB treatment at different levels of BMI change by M/XDR treatment initiation period.

**Supplemental Table B.**
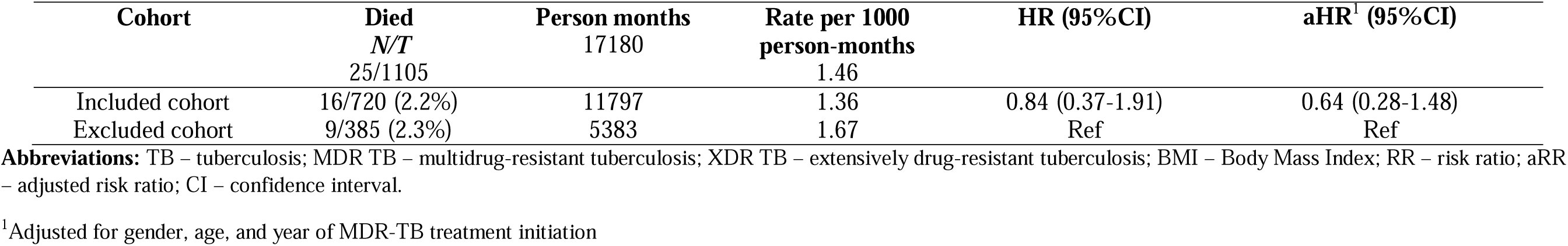
Difference in all-cause mortality during TB treatment among adult M/XDR TB patients included in and excluded from the study, Georgia, 2009-2020 (N=1105) **Purpose:** To assess the difference in hazard of death between included and excluded cohorts to ensure that the study findings are representative of the entire population and not biased due to missing data.

**Supplemental Figure A.**
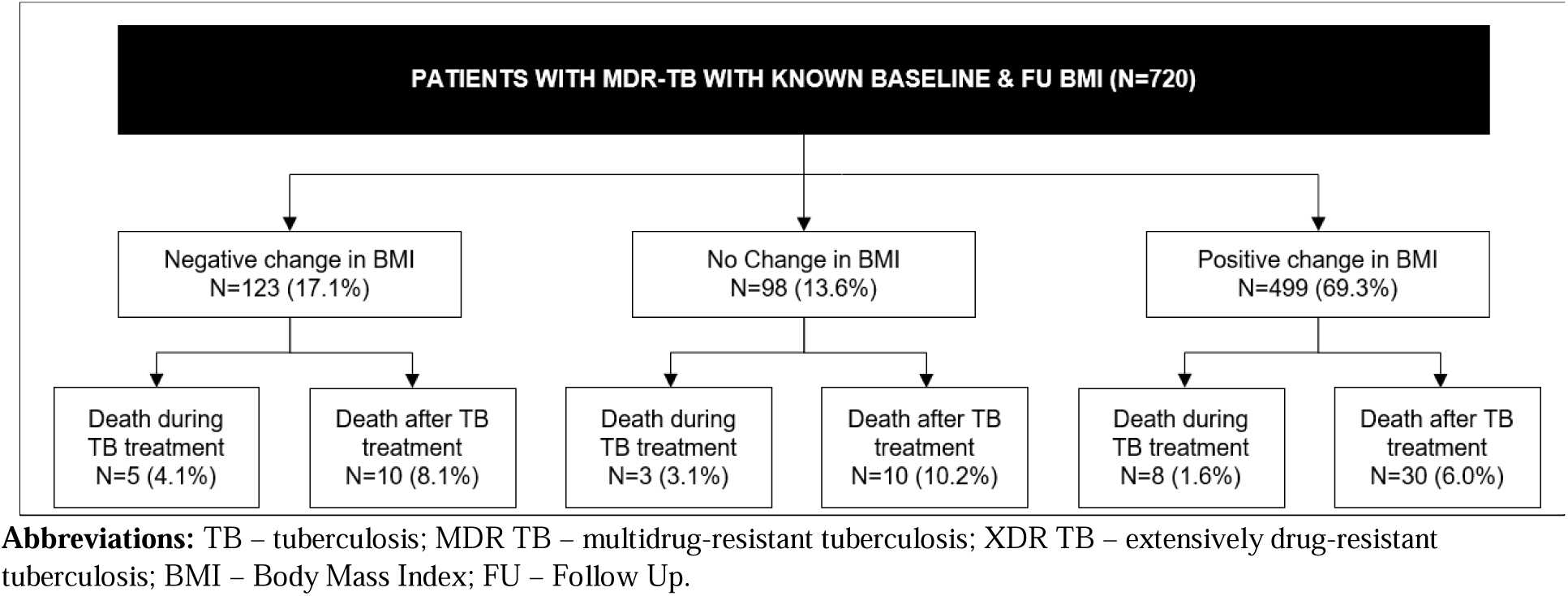
Death during and after TB treatment by baseline BMI change, Georgia, 2009-2020. **Purpose:** To present the numbers of death during and after TB treatment by the change in BMI during Treatment.

