## Supplement for "Lack of weight gain and increased mortality during and after treatment among adults with drug-resistant tuberculosis in Georgia, 2009-2020"

**SUPPLEMENTARY TABLES AND FIGURES**

**Table S1.** Patient and clinical baseline characteristics among M/XDR TB patients by change in BMI, Georgia, 2009-2020 (N=720)

|  | **Total**  N (%)  720 | **Negative change in BMI**  N (%)  123 (17.1%) | **No change in BMI**  N (%)  98 (13.6%) | **Positive change in BMI**  N (%)  499 (69.3%) | **p-value^1^** | **p-value^2^** |
| --- | --- | --- | --- | --- | --- | --- |
| **Baseline BMI (kg/m^2^)** |  |  |  |  | 0.0766^3^ | 0.7870^3^ |
| Median (IQR) | 20.6 (18.8-23.0) | 20.8 (18.9-24.6) | 20.9 (18.9-22.8) | 20.5 (18.8-22.7) |  |  |
| **Categorical Baseline BMI (kg/m^2^)** |  |  |  |  | **0.0011**^4^ | 0.7633^4^ |
| Low (<18.5) | 149 (20.7) | 24 (19.5) | 22 (22.5) | 103 (20.7) |  |  |
| Normal (18.5 – 24.9) | 479 (66.5) | 71 (57.7) | 64 (65.3) | 344 (68.9) |  |  |
| Overweight/Obese (≥ 25) | 92 (12.8) | 28 (22.8) | 12 (12.2) | 52 (10.4) |  |  |
| **Age (years)** |  |  |  |  | 0.1358^3^ | 0.2820^3^ |
| Median (IQR) | 35.5 (26.5-49.0) | 38.0 (28.0-53.0) | 33.5 (26.0-45.0) | 35.0 (26.0-49.0) |  |  |
| **Baseline FBG (mmol/L)** |  |  |  |  | 0.8876^3^ | 0.5244^3^ |
| Median (IQR) |  | 4.9 (4.2-5.7) | 4.9 (4.2-5.5) | 4.9 (4.2-5.7) |  |  |
| Missing^6^ | 58 | 10 | 15 | 33 |  |  |
| **Gender** |  |  |  |  | 0.4041^4^ | 0.6386^4^ |
| Male | 495 (68.8) | 81 (65.9) | 66 (67.3) | 348 (69.7) |  |  |
| Female | 225 (31.2 | 42 (34.1) | 32 (32.7) | 151 (30.3) |  |  |
| **Smoking** |  |  |  |  | 0.5442^4^ | 0.7103^4^ |
| Yes | 281(41.9) | 46 (39.7) | 37 (40.7) | 198 (42.8) |  |  |
| No | 389 (58.1) | 70 (60.3) | 54 (59.3) | 265 (57.2) |  |  |
| Missing^6^ | 50 | 7 | 7 | 36 |  |  |
| **HIV co-infection** |  |  |  |  | 0.6020^5^ | 0.7042^5^ |
| Yes | 17 (2.4) | 2 (1.6) | 3 (3.1) | 12 (2.4) |  |  |
| No | 703 (97.6) | 121 (98.4) | 95 (96.9) | 487 (97.6) |  |  |
| **Hepatitis C co-infection** |  |  |  |  | 0.1125^5^ | 0.4994^5^ |
| Yes | 25 (3.5) | 7 (5.7) | 4 (4.1) | 14 (2.8) |  |  |
| No | 695 (96.5) | 116 (94.3) | 94 (96.0) | 485 (97.2) |  |  |
| **Drug-resistant Type** |  |  |  |  | --- | --- |
| RR | 351 (53.3) | 53 (47.8) | 47 (52.8) | 251 (54.7) |  |  |
| RR & Resistant to FQLs | 77 (11.7) | 19 (17.1) | 10 (11.2) | 48 (10.5) |  |  |
| RR & Resistance to FQL& AG and/or newer anti TB drugs (BDQ, DLM, LZD0 | 231 (35.0) | 39 (35.1) | 32 (36.0) | 160 (34.8) |  |  |
| Missing^6^ | 62 | 13 | 9 | 40 |  |  |
| **Baseline sputum smear** |  |  |  |  | 0.5626^4^ | 0.9546^4^ |
| Positive | 455 (63.5) | 74 (61.2) | 63 (64.3) | 318 (64.0) |  |  |
| Negative | 261 (36.5) | 47 (38.8) | 35 (35.7) | 179 (36.0) |  |  |
| Missing^6^ | 4 | 2 | 0 | 2 |  |  |
| **CXR cavitation** |  |  |  |  | 0.1031^4^ | 0.9826^4^ |
| Yes | 240 (33.5) | 33 (27.0) | 34 (34.7) | 173 (34.8) |  |  |
| No | 477 (66.5) | 89 (73.0) | 64 (65.3) | 324 (66.2) |  |  |
| Missing^6^ | 3 | 1 | 0 | 2 |  |  |
| **Treatment period** |  |  |  |  | 0.7088^4^ | 0.3374^4^ |
| 2009-2015 cohort | 391 (54.3) | 64 (52.0) | 58 (59.2) | 269 (53.9) |  |  |
| 2016-2019 cohort | 329 (45.7) | 59 (48.0) | 40 (40.8) | 230 (46.1) |  |  |

**Abbreviations:** TB – tuberculosis; MDR TB – multidrug-resistant tuberculosis; XDR TB – extensively drug-resistant tuberculosis; BMI – Body Mass Index; IQR – interquartile range; FBG – Fasting Blood Glucose; HIV – human immunodeficiency virus; RR – Rifampicin Resistance; AG – Aminoglycosides; FQL – fluoroquinolones; BDQ – Bedaquiline; DLM – Delamanid; LZD – Linezolid

^1^p-value comparing values for patients with negative BMI relative change versus patients with positive BMI relative change.

^2^p-value comparing values for patients with no BMI relative change versus patients with positive BMI relative change.

^3^Wilcoxon rank sum two-sample test was used (for non-normally distributed continuous variables)

^4^Chi-square test was used for categorical variables.

^5^Fisher exact test was used for categorical variables when the expected cell size was less than 5.

^6^Missing is not used as a group and is not included in the analysis.

**Table S2.** Associations between change in BMI with all-cause mortality during TB treatment stratified by period of MDR-TB treatment initiation, among adult M/XDR TB patients, Georgia, 2009-2020 (N=720)

| **Baseline BMI Status** | **BMI percentage change** | **Died**  ***N/T***  16/720 | **Person months**  11797 | **Rate per 1000 person-months** 1.36 | **HR (95%CI)** | **aHR**^1^ **(95%CI)** | **aHR**^2^ **(95%CI)** |
| --- | --- | --- | --- | --- | --- | --- | --- |
| **Total cohort**  n=720 | Negative | 5/123 (4.1%) | 1900 | 2.63 | 2.77 (0.91-8.49) | 2.70 (0.87-8.34) | 2.99 (0.96-9.34) |
|  | No | 3/98 (3.1%) | 1705 | 1.76 | 1.85 (0.49-6.98) | 2.09 (0.55-7.96) | 2.08 (0.55-7.96) |
|  | Positive | 8/499 (1.6%) | 8192 | 0.98 | Ref | Ref | Ref |
| 2009-2015 cohort  n=391 | Negative | 2/64 (3.1%) | 1134 | 1.76 | 2.22 (0.41-12.17) | 2.01 (0.36-11.09) | 2.65 (0.45-15.47) |
|  | No | 2/58 (3.5%) | 1097 | 1.82 | 2.30 (0.42-12.59) | 2.62 (0.47-14.64) | 2.58 (0.43-15.45) |
|  | Positive | 4/269 (1.5%) | 4818 | 0.83 | Ref | Ref | Ref |
| 2016-2020 cohort  n=329 | Negative | 3/59 (5.1%) | 766 | 3.92 | 3.38 (0.76-15.10) | 2.93 (0.65-13.27) | 2.99 (0.65-13.68) |
|  | No | 1/40 (2.5%) | 608 | 1.65 | 1.35 (0.15-12.10) | 1.81 (0.20-16.86) | 1.95 (0.20-18.95) |
|  | Positive | 4/230 (1.7%) | 3374 | 1.19 | Ref | Ref | Ref |

**Abbreviations:** TB – tuberculosis; MDR TB – multidrug-resistant tuberculosis; XDR TB – extensively drug-resistant tuberculosis; BMI – Body Mass Index; HR – hazard ratio; aHR – adjusted hazard ratio; CI – confidence interval.

^1^Adjusted for gender, age, and year of MDR-TB treatment initiation

^2^Adjusted for gender, age, year of MDR-TB treatment initiation, and baseline sputum smear results.

**Table S3.** Difference in all-cause mortality during TB treatment among adult M/XDR TB patients included in and excluded from the study, Georgia, 2009-2020 (N=1105)

| **Cohort** | **Died**  ***N/T***  25/1105 | **Person months**  17180 | **Rate per 1000 person-months** 1.46 | **HR (95%CI)** | **aHR**^1^ **(95%CI)** |
| --- | --- | --- | --- | --- | --- |
| Included cohort | 16/720 (2.2%) | 11797 | 1.36 | 0.84 (0.37-1.91) | 0.64 (0.28-1.48) |
| Excluded cohort | 9/385 (2.3%) | 5383 | 1.67 | Ref | Ref |

**Abbreviations:** TB – tuberculosis; MDR TB – multidrug-resistant tuberculosis; XDR TB – extensively drug-resistant tuberculosis; BMI – Body Mass Index; RR – risk ratio; aRR – adjusted risk ratio; CI – confidence interval.

^1^Adjusted for gender, age, and year of MDR-TB treatment initiation
